# Germline sequencing of DNA-damage-repair genes in two hereditary prostate cancer cohorts reveals new disease risk-associated gene variants

**DOI:** 10.1101/2022.04.11.22273677

**Authors:** Georgea R. Foley, James R. Marthick, Sionne E. Lucas, Kelsie Raspin, Annette Banks, Janet L. Stanford, Elaine A. Ostrander, Liesel M. FitzGerald, Joanne L. Dickinson

## Abstract

**Background:** Knowledge of rare, inherited variants in DNA damage repair (DDR) genes is informing clinical management in common cancers. However, defining the rare disease-associated variants in prostate cancer (PrCa) is challenging due to their low frequency.

**Method:** Here, whole-genome and -exome sequencing data from two independent, high-risk Australian and North American familial PrCa datasets were interrogated for novel, rare DDR variants. Segregating, high-risk, likely pathogenic DDR gene variants were identified and subsequently genotyped in 1,963 individuals (700 familial and 459 sporadic PrCa cases, 482 unaffected relatives, and 322 screened controls) and association analyses performed accounting for relatedness (M_QLS_).

**Results:** Rare variants significantly associated with PrCa risk were identified in *ERCC3* (rs145201970, p=2.57×10^−4^) and *BRIP1* (rs4988345, p=0.025) in the combined datasets. A *PARP2* (rs200603922, p=0.028) variant in the Australian dataset and a *MUTYH* (rs36053993, p=0.031) variant in the North American dataset were also associated with PrCa risk. No evidence for a younger age or higher-grade disease at diagnosis was evident in variant carriers.

**Conclusions:** Here, we provide new evidence for four novel germline DDR PrCa risk variants. Defining the full spectrum of PrCa associated DDR genes is important for effective clinical screening and disease management.

## BACKGROUND

Prostate cancer (PrCa) is responsible for a significant proportion of cancer-related deaths in men worldwide and presents a substantial health burden in terms of morbidity, mental health, and economic costs associated with treatment. A considerable percentage of men with advanced disease harbour clinically actionable variants, many of which are aberrations in DNA damage repair (DDR) genes (1, 2, 3, 4, 5, 6). Notably, germline variants in these genes have been observed in 8–16% of metastatic PrCa patients (1, 4, 7).

Despite recognition of their potential, there remains a significant gap in our understanding of the spectrum of DDR gene variants contributing to PrCa risk (7, 8, 9, 10, 11). Variants in several of these genes, particularly *ATM* and *BRCA1/2*, have been associated with a poorer prognosis, differing responses to treatment, and more aggressive disease (12, 13, 14, 15, 16). Importantly, tumours harbouring loss-of-function mutations in DDR genes exhibit a therapeutic response to poly (ADP-ribose) polymerase inhibitors (PARPi)(17) and platinum-based chemotherapy (18). Thus, screening for clinically actionable germline variants in PrCa patients, particularly those with a significant family history together with advanced disease, represents an important strategy to improve PrCa outcomes. The rarity of these variants in population-based PrCa datasets, which represent the majority of PrCa DDR gene discovery studies to date, has hampered research efforts. In addition, many of these studies have not differentiated between germline and acquired mutations, and those variants that have been identified remain largely of unknown clinical significance.

Curation of the full spectrum of DDR genetic variants contributing to PrCa risk has significant potential in the healthcare setting, where precision medicine can be implemented for both diagnosis and treatment. Moreover, the observation that germline and acquired mutations are frequently identified in the same DDR genes underscores the importance of these pathways in tumour development. Here, we interrogated whole-genome and -exome germline data from two high-risk familial PrCa datasets with the aim of identifying novel, rare DDR gene variants contributing to PrCa risk.

## METHODS

The aim of this study was to identify novel rare putative pathogenic variants in DDR genes using two large familial prostate cancer genetic cohorts from Australia and North America.

### Study Resources

This study utilises clinicopathological and genetic data available from two independent PrCa resources: the Australian resource, consisting of the Tasmanian Familial Prostate Cancer Study and the population-based Tasmanian Prostate Cancer Case-Control Study, and the North American resource, consisting of the Prostate Cancer Genetic Research Study (*PROGRESS*) from the Fred Hutchinson Cancer Center (FHCC).

The Tasmanian Familial Prostate Cancer Study included 73 PrCa families comprising DNA from 379 affected men and 471 unaffected male and female relatives of Northern European heritage (19, 20, 21). The study was initiated in Tasmania in the late 1990s, prior to the implementation of wide-spread prostate-specific antigen (PSA) testing as a PrCa screening tool. Families with more than two affected first-degree relatives spanning two or more generations are included and were identified through interrogation of the Menzies Genealogical database and the Tasmanian Cancer Registry (TCR), in addition to collaboration with local clinicians.

The second, population-based Tasmanian Prostate Cancer Case-Control Study comprises 459 cases and 322 male controls of Northern European ancestry (19, 20, 21). Cases were identified from the TCR. Controls were selected at random from the Tasmanian electoral roll, frequency matched by five-year age groups to the cases, and checked bi-annually against the TCR for PrCa diagnosis.

The FHCC resource comprises *PROGRESS*, which includes >300 PrCa families from across North America (22). Whole exome sequencing (WES) data were available for 130 families, which included 11 older unaffected men and 321 affected men. Men prioritised for WES were those diagnosed with an early-onset/aggressive disease phenotype, uncle-nephew and/or cousin pairs from families with densely aggregated affected men.

Details of the clinical data available for these resources are presented in Table 1.

**Table 1:** Clinical characteristics of study resources with available genetic data

|  | **Australian Familial**<br>**Cases, n (%)** | **Australian Sporadic**<br>**Cases, n (%)** | ***PROGRESS* Familial**<br>**Cases, n (%)** |
| --- | --- | --- | --- |
| Age at Diagnosis |  |  |  |
| <60 | 93 (24.54%) | 137 (29.85%) | 108 (33.64%) |
| 60-64 | 98 (25.86%) | 170 (37.04%) | 75 (23.36%) |
| 65-69 | 97 (20.32%) | 125 (27.23%) | 77 (23.99%) |
| ≥70 | 107 (28.23%) | 26 (5.66%) | 61 (19.00%) |
| Missing | 4 (1.06%) | 1 (0.22%) | n.a. |
| Age at Diagnosis,<br>Median (IQR) | 64.82 (60.06-71.50) | 62.59 (59.26-66.05) | 63 (57.0-68.0) |
| Years between<br>Diagnosis and Death |  |  |  |
| <5 | 19 (5.01%) | 25 (5.45%) | 11 (3.43%) |
| 5-9 | 39 (10.29%) | 54 (11.76%) | 36 (11.21%) |
| 10-19 | 81 (21.37%) | 103 (22.44%) | 65 (20.25%) |
| ≥20 | 18 (4.75%) | 19 (4.14%) | 7 (2.18%) |
| Missing | 2 (0.53%) | n.a. | n.a. |
| n.a. | 220 (58.05%) | 258 (56.21%) | 202 (62.93%) |
| Years between<br>Diagnosis and Death,<br>Median (IQR) | 11.62 (8.75-16.10) | 11.82 (7.64-15.52) | 11.00 (6.50-15.0) |
| Cause of Death |  |  |  |
| PrCa | 49 (12.93%) | 68 (14.81%) | 41 (12.77%) |
| Other | 91 (24.01%) | 133 (28.98%) | 72 (22.43%) |
| Not Processed | 2 (0.53%) | n.a. | n.a. |
| Missing | 17 (4.49%) | 1 (0.22%) | 6 (1.87%) |
| n.a. | 220 (58.05%) | 257 (55.99%) | 202 (62.93%) |
| **Total** | **379** | **459** | **321** |

IQR = Interquartile Range; n.a. = Not Applicable; PrCa = Prostate Cancer

Additionally, age-at-diagnosis and Gleason score (GS) data from 2,126 participants enrolled in the population-based Prostate Cancer Outcomes Registry - Tasmania (PCOR-TAS) were available for clinicopathological analyses. PCOR-TAS was established in 2015, with an aim to improve all aspects of the quality of care for men diagnosed with PrCa. The opt-out registry is an ongoing initiative that records data on the diagnosis, treatment, outcomes, and quality of life for all Tasmanians diagnosed with PrCa. Details of the registry, including data collection methods, have been described previously (23). As of the 15^th^ of July 2021, 2,126 men had been recruited into PCOR-TAS, with ∼3% having opted out of the registry.

### Whole-Genome Sequencing and Bioinformatic Sequence Analysis

Whole-genome sequencing (WGS) data were generated from germline DNA (additional details: Supplementary Method 1) for 54 individuals from eight Australian families (Supplementary Table S1) and seven unaffected men from the Australian Case-Control Study.

Of the familial individuals, 43 had been diagnosed with PrCa, with the remaining individuals comprising a female relative with a self-reported breast cancer diagnosis (n=1) and unaffected male relatives (n=10). WGS (mean coverage = 38.7x; range = 29.2x-49.8x) was completed in five instalments at the Australian Genome Research Facility (Melbourne, Australia), the Ramaciotti Centre for Genomics (Sydney, Australia), and the Texas Biomedical Research Institute (San Antonio, Texas). Sequence data were aligned to the hg19 reference genome with BWA-MEM (24) and variants were called with GATK (25), using bcbio (26).

### Variant Filtering, Prioritisation, and Validation

A panel of 35 genes involved in DDR pathways was compiled (Table 2), in addition to the established PrCa risk gene, *HOXB13* (27). Variants located in a genomic window 1000bp up and downstream of the nominated candidate genes were extracted from WGS data using bcftools (28) and annotated using ANNOVAR (29). Included genes and genomic positions can be found in Supplementary Table S2.

**Table 2:**
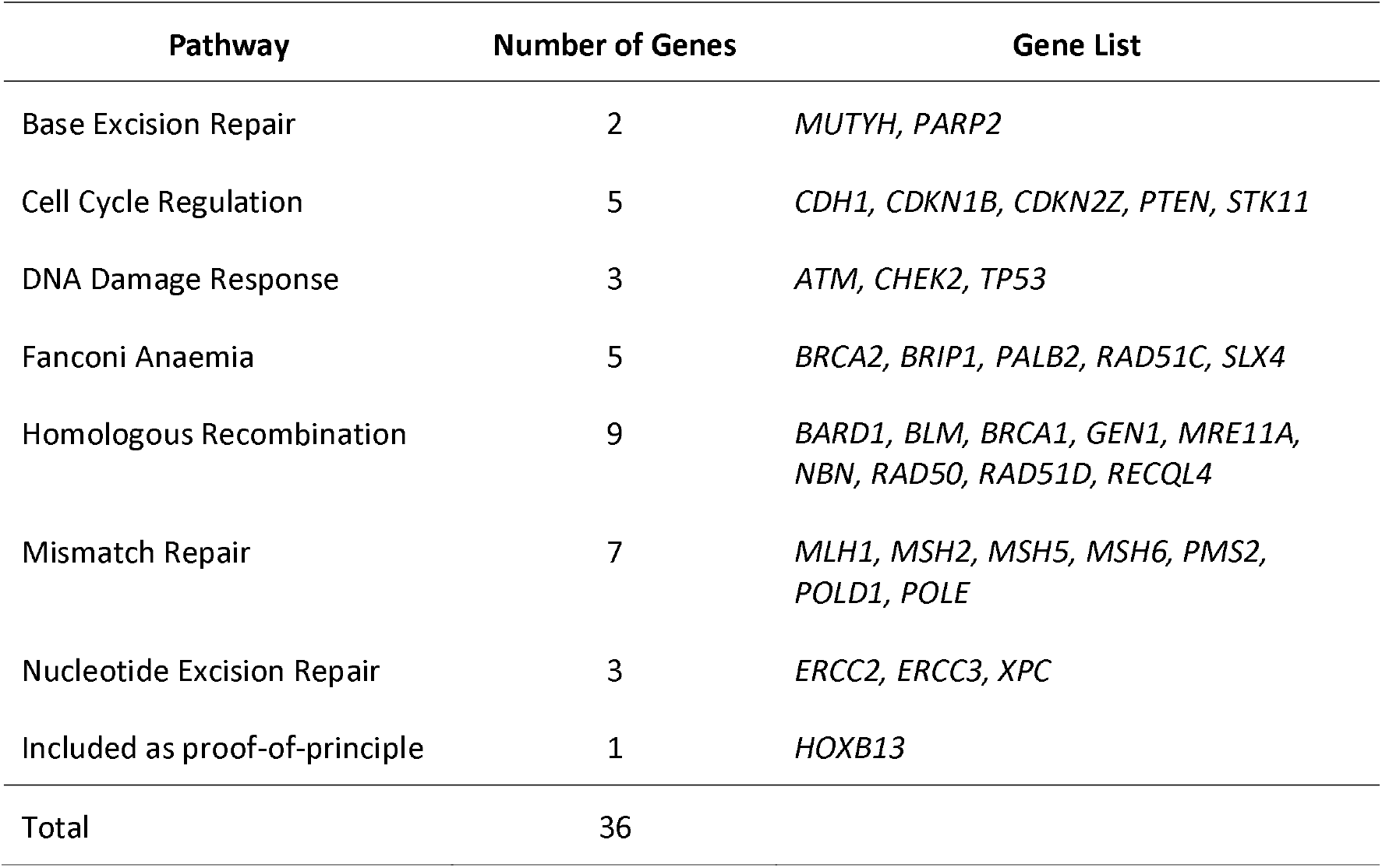
DNA repair pathway genes included for analysis

| Pathway | Number of Genes | Gene List |
| --- | --- | --- |
| Base Excision Repair | 2 | *MUTYH, PARP2* |
| Cell Cycle Regulation | 5 | *CDH1, CDKN1B, CDKN2Z, PTEN, STK11* |
| DNA Damage Response | 3 | *ATM, CHEK2, TP53* |
| Fanconi Anaemia | 5 | *BRCA2, BRIP1, PALB2, RAD51C, SLX4* |
| Homologous Recombination | 9 | *BARD1, BLM, BRCA1, GEN1, MRE11A, NBN, RAD50, RAD51D, RECQL4* |
| Mismatch Repair | 7 | *MLH1, MSH2, MSH5, MSH6, PMS2, POLD1, POLE* |
| Nucleotide Excision Repair | 3 | *ERCC2, ERCC3, XPC* |
| Included as proof-of-principle | 1 | *HOXB13* |
| Total | 36 |  |

Variant filtering and prioritisation occurred according to a range of criteria (Figure 1). Variants were filtered to include those with a minor allele frequency (MAF) <1% in gnomAD non-Finnish Europeans (NFE) and Combined Annotation-Dependent Depletion (CADD) score >15, with further prioritisation informed by predicted mutation function (e.g., nonsense > missense > splicing > synonymous). Variants were excluded if present in >1 of the seven screened unaffected male control genomes, or if present only in PrCa unaffected familial individuals.

**Figure 1:**
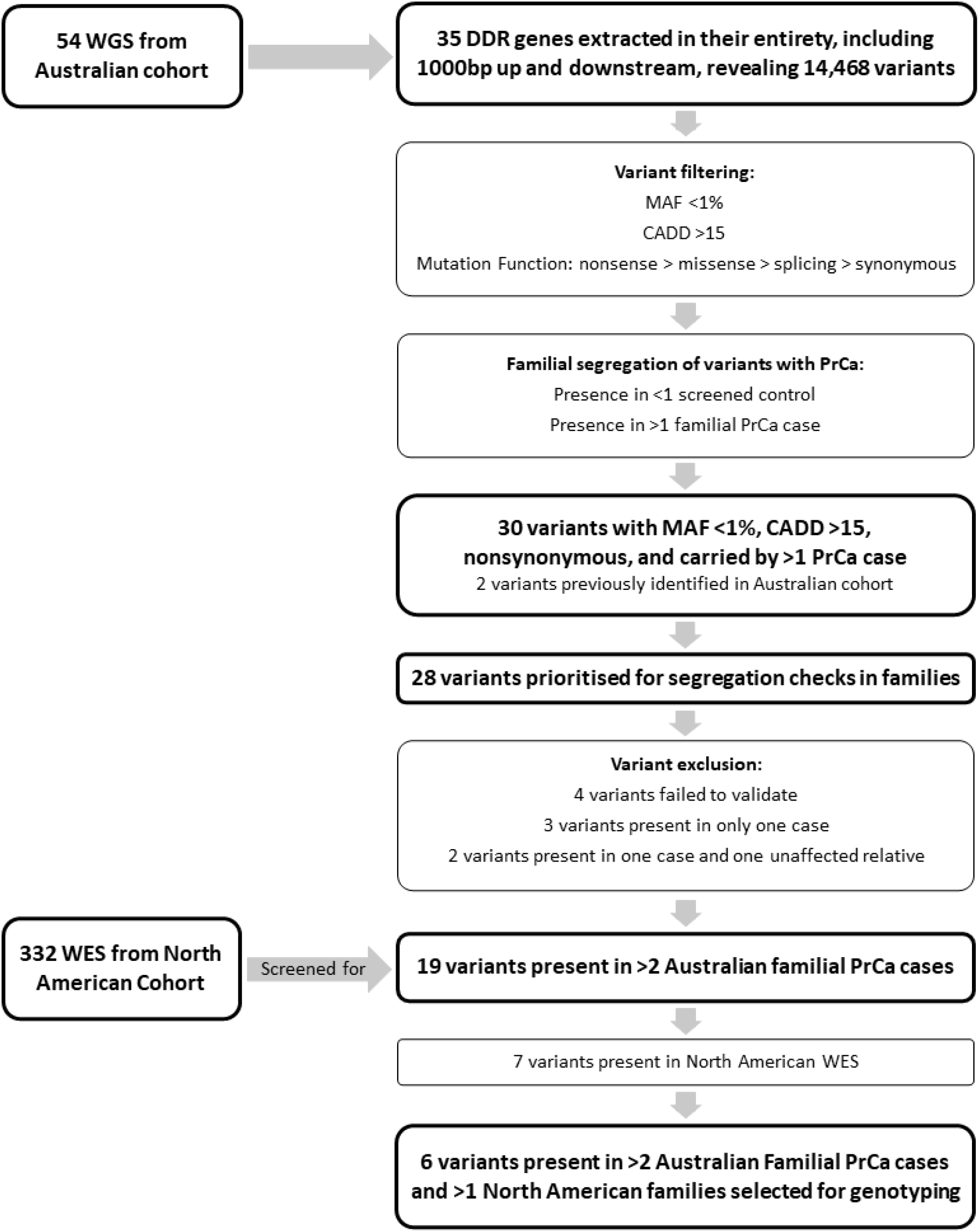
Variant Filtering and Prioritisation Schematic. Flow chart outlining genetic analysis pipeline including variant filtering and prioritisation of variants for follow up.

Short-listed variants (MAF <1%, CADD >15, nonsynonymous, and carried by >1 PrCa case; (Figure 1), which had been validated by Sanger sequencing on the ABI 3500 Genetic Analyser (Applied Biosystems), were genotyped in additional non-WGS relatives to determine segregation in the relevant discovery family. Primers were designed to amplify fragments approximately 300bp in length for each of the selected variants. Primer sequences are presented in Supplementary Table S3, and PCR conditions are available upon request.

### Additional Genotyping in Expanded Australian Resources and Statistical Analysis

Six prioritised variants (Figure 1) were genotyped in the full Australian familial and case-control resources, using TaqMan™ genotyping assays (ThermoFisher Scientific; Supplementary Table S4) on the LightCycler® 480 system (Roche). Existing whole exome data were interrogated for prioritised gene variants in the *PROGRESS* study individuals. Association between genotyped variants and PrCa risk was tested for using Modified Quasi-Likelihood Score (M_QLS_) analysis (30) (additional details: Supplementary Method 2). Population prevalence of PrCa was set at one in seven, and the analyses were conducted in the Australian familial and case-control datasets alone, the FHCC *PROGRESS* cohort alone, and all datasets combined. For DDR variants significantly associated with PrCa risk, diagnosis age of variant carriers was compared with population-based PCOR-TAS cases (TAS) or the full *PROGRESS* cohort (FHCC) and the proportion of variant carriers across several age-at-diagnosis categories (<50, <55, <60, <65, <70) was examined. GS at diagnosis of variant carriers was also recorded where available.

## RESULTS

### Clinical Characteristics of Australian and North American PrCa Resources

Clinical characteristics of the study resources are presented in Table 1. Age-at-diagnosis, time interval between diagnosis and death, and proportion of PrCa-specific deaths were similar across the datasets.

### Identification of Candidate Rare DDR PrCa Risk Variants

WGS data were interrogated for rare, potentially pathogenic variants in 35 DDR genes (Table 2). Initial filtering identified 30 variants in 20 genes, of which two in *HOXB13* and RAD51C have previously been shown to be significantly associated with PrCa risk in our Australian cohort (19, 20), providing proof-of-principle for our approach. Of the 28 remaining variants, four failed to validate via Sanger sequencing and were excluded from further investigation.

Additional non-WGS affected and unaffected relatives with DNA from each of the Australian discovery families underwent Sanger sequencing to determine segregation with disease of the remaining 24 variants (Table 3). Five variants were subsequently excluded: three variants that were each only present in a single affected man and two variants that were only present in a single affected man and one unaffected relative. The remaining 19 variants, *ATM* rs56128736, *BARD1* rs3738888, *BRCA1* rs28897673, *BRCA2* rs28897727, *BRCA2* rs55639415, *BRCA2* rs786202915, *BRIP1* rs4988345, *ERCC2* rs142568756, *ERCC3* rs145201970, *MRE11* rs777373591, *MSH6* rs142254875, *MUTYH* rs36053993, *PARP2* rs200603922, *PMS2* rs1554304601, *POLE* chr12: 133219216, *POLE* rs41561818, *PTEN* rs587779989, *PTEN* rs773513402, and *RECQL4* rs780723602, were present in at least two affected relatives from the Australian discovery cohort.

**Table 3:** Putative pathogenic mutations identified in DDR genes in Australian discovery and North American families

| Gene | Variant | Amino Acid Change | MAF (gnomAD NFE) | CADD | Australian Discovery Familial Cohort |  |  | North American Familial Cohort |  |  |
| --- | --- | --- | --- | --- | --- | --- | --- | --- | --- | --- |
|  |  |  |  |  | PrCa Affected Carriers | Total Carriers | Number of Families | PrCa Affected Carriers | Total Carriers | Number of Families |
| *ATM* | rs55801750 | C2464R | 9E-04 | 22.7 | 1 | 2 | 1 | - | - | - |
| *ATM* | rs55982963 | R2719H | 1E-04 | 29.3 | 1 | 2 | 1 | - | - | - |
| *ATM* | rs56128736 | V410A | 0.0021 | 23.1 | 3 | 6 | 2 | 2 | 2 | 1 |
| *ATM* | rs767507047 | Y2954C | 6.48E-05 | 28.6 | 1 | 1 | 1 | - | - | - |
| *BRCA1* | rs28897673 | Y58C | 1E-04 | 23.6 | 2 | 4 | 1 | - | - | - |
| *BRCA2* | rs55639415 | S1733F | 4.71E-05 | 15.43 | 2 | 4 | 1 | - | - | - |
| *BRCA2* | rs56403624 | E462G | 4.29E-04 | 19.02 | 1 | 1 | 1 | - | - | - |
| *BRCA2* | rs786202915 | F2254Y | n.a. | 16.65 | 3 | 8 | 1 | - | - | - |
| *ERCC2* | rs142568756 | V536M | 0.0005 | 29.1 | 2 | 4 | 1 | - | - | - |
| *MRE11* | rs777373591 | P132S | 1.77E-05 | 27.2 | 2 | 4 | 1 | - | - | - |
| *MSH6* | rs142254875 | P943S | 0.0001 | 22.2 | 2 | 6 | 1 | - | - | - |
| *PMS2* | rs1554304601 | A116V | n.a. | 31 | 3 | 7 | 1 | - | - | - |
| *POLE* | chr12: 133219216 | P1610A | n.a. | 22.8 | 3 | 9 | 1 | - | - | - |
| *POLE* | rs36120395 | P697R | 0.0016 | 20.3 | 1 | 1 | 1 | - | - | - |
| *POLE* | rs41561818 | A1420V | 0.0044 | 21.2 | 2 | 3 | 1 | - | - | - |
| *PTEN* | rs587779989 | n.a. | n.a. | 19.81 | 3 | 6 | 1 | - | - | - |
| *PTEN* | rs773513402 | n.a. | 0.0003 | 15.18 | 2 | 5 | 1 | - | - | - |
| *RECQL4* | rs780723602 | I920V | 9E-06 | 22.3 | 4 | 11 | 1 | - | - | - |
| *BARD1* | rs3738888 | R188C | 0.0063 | 26.4 | 2 | 5 | 1 | 4 | 4 | 4 |
| *BRCA2* | rs28897727 | D1420Y | 0.0098 | 15.85 | 3 | 9 | 1 | 4 | 5 | 3 |
| *BRIP1* | rs4988345 | R173C | 0.0043 | 25.5 | 2 | 5 | 1 | 6 | 6 | 6 |
| *ERCC3* | rs145201970 | R283C | 0.0017 | 26.7 | 2 | 7 | 1 | 5 | 5 | 3 |
| *MUTYH* | rs36053993 | G368D | 0.0047 | 32 | 2 | 5 | 2 | 9 | 9 | 4 |
| *PARP2* | rs200603922 | R15G | 0.0012 | 15.32 | 4 | 6 | 1 | 2 | 2 | 2 |

n.a. = Not Applicable

For further prioritisation, we then sought to determine whether any of the 19 variants were present in the North American *PROGRESS* families. Examination of exome data from 332 individuals revealed seven variants in 34 cases from 22 kindreds. Four variants, *ATM* rs56128736, *BRCA2* rs28897727, *ERCC3* rs145201970, and *MUTYH* rs36053993, were present in two or more PrCa cases in a single family (Table 3).

Six DDR variants, *BARD1* rs3738888, *BRCA2* rs28897727, *BRIP1* rs4988345, *ERCC3* rs145201970, *MUTYH* rs36053993, and *PARP2* rs200603922, that segregated with disease in an Australian PrCa family and were present in two or more *PROGRESS* families, were selected for additional investigation (Table 3). These variants were genotyped in the extended Australian familial and case-control resources via TaqMan genotyping. All six variants were identified in additional individuals (n_range_=9 to 33; Supplementary Table S5) within the Australian datasets, and all except *MUTYH* rs36053993 were observed in additional familial PrCa cases. With the inclusion of the *PROGRESS* dataset, the *BARD1* rs3738888 and *BRIP1* rs4988345 variants were each observed in the most PrCa cases (n=22), which included ten and nine sporadic cases, respectively. The predicted pathogenicity of these variants was determined using multiple bioinformatic tools (additional details: Supplementary Method 3) and outputs are shown in Table 4.

**Table 4:** Predicted pathogenicity of prioritised DDR gene variants

| Gene | Variant | Allele Change | AA Change | CADD | DANN* | SIFT | PROVEAN | PolyPhen | Mutation Taster (Rank Score) | Mutation Assessor | FATHMM† (Coding) |
| --- | --- | --- | --- | --- | --- | --- | --- | --- | --- | --- | --- |
| *BARD1* | rs3738888 | G>A | R658C | 26.4 | 0.999 | 0.008 (D) | -4.02 (De) | 1 (P) | 0.462 (D) | 2.12 (M) | 0.9778 |
| *BRCA2* | rs28897727 | G>T | D1420Y | 15.85 | 0.988 | 0.030 (D) | -6.60 (De) | 0.03 (B) | 0.09 (N) | 2.15 (M) | 0.49798 |
| *BRIP1* | rs4988345 | G>A | R173C | 25.5 | 0.999 | 0.001 (D) | -2.54 (De) | 1 (P) | 0.81 (D) | 2.67 (M) | 0.93639 |
| *ERCC3* | rs145201970 | G>A | R283C | 26.7 | 0.999 | 0.000 (D) | -7.59 (De) | 0.995 (P) | 0.81 (D) | 3.31 (M) | 0.99364 |
| *MUTYH* | rs36053993 | C>T | G368D | 32 | 0.998 | 0.000 (D) | -6.46 (De) | 1 (P) | 0.81 (D) | 4.09 (H) | 0.99757 |
| *PARP2* | rs200603922 | A>G | R15G | 15.32 | 0.8 | 0.153 (T) | -1.04 (N) | 0 (B) | 0.09 (N) | 0.695 (N) | 0.00048 |

D = Damaging; T = Tolerated; De = Deleterious; N = Neutral; P = Probably Damaging; B = Benign; M = Medium; H = High. \*DANN predictions use a scoring system between 0 and 1, with scores closer to one indicating greater predicted pathogenicity. †FATHMM predictions use a scoring system between 0 and 1, with scores closer to one indicating greater predicted pathogenicity.

### Statistical Analysis

Genotypes were available for six variants in 1,963 individuals, including 700 familial and 459 sporadic PrCa cases overall. M_QLS_ association analysis permitted the inclusion of related and unrelated individuals while also appropriately controlling for Type 1 error (30). In the Australian dataset, a significant association was observed between *PARP2* rs200603922 and PrCa risk (p=0.028), whilst in the *PROGRESS* dataset, a significant association was observed between *BRIP1* rs4988345 (p=0.034), *ERCC3* rs145201970 (p=0.010), and *MUTYH* rs36053993 (p=0.031) and PrCa risk (Table 5). In the combined Australian and *PROGRESS* datasets, a significant association with PrCa risk was observed between *BRIP1* rs4988345 (p=0.025) and *ERCC3* rs145201970 (p=2.57×10^−4^). The *ERCC3* variant remains significant following Bonferroni correction for multiple testing. PrCa status of variant carriers is provided in Supplementary Table S5 and clinical characteristics of affected familial carriers are available upon request.

**Table 5:** Carrier frequency and statistical analysis of variants

| Gene | Variant | Australian Familial and Sporadic Prostate Cancer |  |  | North American Familial *PROGRESS* Cohort |  |  | Tasmanian Familial Prostate Cancer Study & *PROGRESS* |  |  |
| --- | --- | --- | --- | --- | --- | --- | --- | --- | --- | --- |
|  |  | Total Carriers<br>(% Cases)* | M<sub>QLS</sub><br>P-Value | M<sub>QLS</sub><br>Odds Ratio | Total Carriers<br>(% Cases)* | M<sub>QLS</sub><br>P-Value | M<sub>QLS</sub><br>Odds Ratio | Total Carriers<br>(% Cases)* | M<sub>QLS</sub><br>P-Value | M<sub>QLS</sub><br>Odds Ratio |
| *BARD1* | rs3738888 | 31 (58.1%) | 0.407 | 1.7 | 4 (100%) | 0.318 | n.a. | 35 (62.9%) | 0.066 | 1.9 |
| *BRCA2* | rs28897727 | 24 (54.2%) | 0.063 | n.a. | 5 (80%) | 0.157 | n.a. | 29 (58.6%) | 0.193 | n.a. |
| *BRIP1* | rs4988345 | 25 (64.0%) | 0.118 | 3.1 | 6 (100%) | **0.034** | **n.a.** | 31 (71.0%) | **0.025** | **3.1** |
| *ERCC3* | rs145201970 | 16 (50.0%) | 0.554 | 1 | 5 (100%) | **0.010** | **n.a.** | 21 (61.9%) | **2.57x10<sup>-4</sup>** | **1.7** |
| *MUTYH* | rs36053993 | 23 (26.1%) | 0.630 | 0.4 | 9 (100%) | **0.031** | **n.a.** | 32 (46.9%) | 0.201 | 0.8 |
| *PARP2* | rs200603922 | 14 (71.4%) | **0.028** | **n.a.** | 2 (100%) | 0.388 | n.a. | 16 (75.0%) | 0.162 | n.a. |

n.a. = Not Applicable, as the odds ratio cannot be calculated when no carriers in controls are identified. \*% of variant carriers that are cases.

Age-at-diagnosis amongst variant carriers was compared to relevant population datasets (Figure 2) with a shift towards younger age-at-diagnosis observed, most evident in Australian sporadic carriers compared with the population-based PCOR-TAS cohort. While a slightly higher proportion of DDR variant carriers was observed in cases diagnosed before 55 years of age (9.4%; Table 6), variant carriers were relatively consistent at ∼6% across the remaining age at diagnosis categories (Table 6). Population data from PCOR-TAS reveals that ∼20% of Tasmanian men are diagnosed with a GS≥8. Of men carrying a risk DDR variant, 23% (15/66) were diagnosed with GS≥8, while 58% (38/66) of rare variant carriers were diagnosed with a GS≤6.

**Table 6:** DDR variant rates by age at diagnosis

|  | PrCa cases meeting criteria<br>N (%) | Variant carriers meeting criteria*<br>N (%) |
| --- | --- | --- |
| **Total PrCa Cases** | 1159 | 81 |
| **Age at Diagnosis <50 years** |  |  |
| All PrCa cases | 29 2.50% | 2 6.90% |
| DDR variant carriers | 2 2.47% |  |
| **Age at Diagnosis <55 years** |  |  |
| All PrCa cases | 117 10.09% | 11 9.40% |
| DDR variant carriers | 11 13.58% |  |
| **Age at Diagnosis <60 years** |  |  |
| All PrCa cases | 344 29.68% | 20 5.81% |
| DDR variant carriers | 20 24.69% |  |
| **Age at Diagnosis <65 years** |  |  |
| All PrCa cases | 693 59.79% | 38 5.48% |
| DDR variant carriers | 38 46.91% |  |
| **Age at Diagnosis <70 years** |  |  |
| All PrCa cases | 975 68.59% | 62 6.36% |
| DDR variant carriers | 62 76.54% |  |

\*Values represent the number and percentage of Tasmanian and North American cases diagnosed within the specified age bracket who carry a rare DDR risk variant.

**Figure 2:**
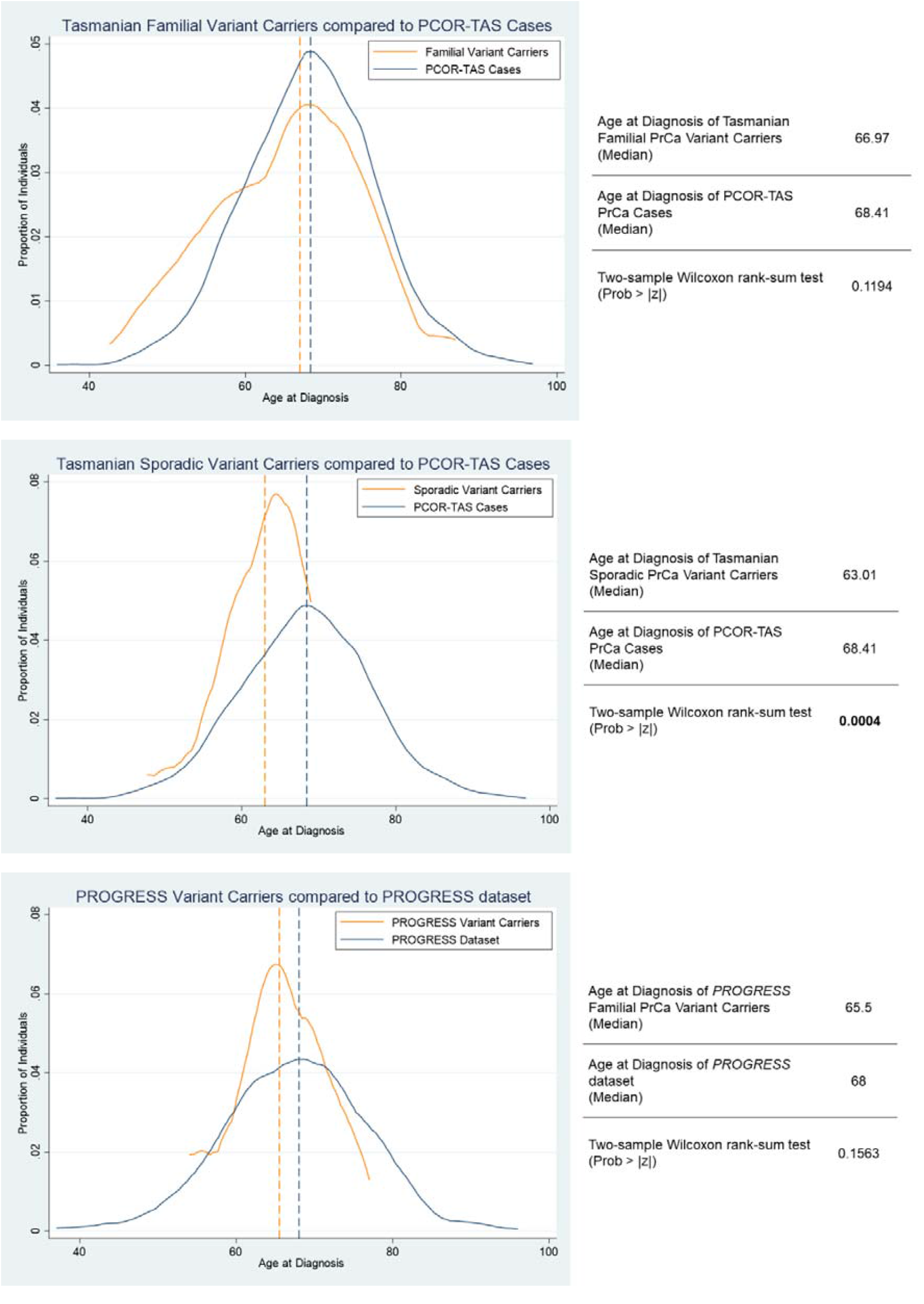
Age at diagnosis of DDR gene variant carriers** compared to available population-based datasets. Age at diagnosis of variant carriers compared with a comparable population dataset (presented as proportion of individuals versus age at diagnosis). Panel A -familial prostate cancer variant carriers (n=31) overlayed with unscreened PCOR-Tas case population (n=2126); panel B- sporadic prostate cancer variant carriers (n=22) compared with unscreened PCOR-Tas population; and panel C – *PROGRESS* dataset variant carriers (n=28) compared with non-variant carriers from the *PROGRESS* dataset. **DDR variant carriers = those carrying a pathogenic variant in *ERCC3, BRIP1, PARP2, MUTYH*, RAD51C and BRCA2.

## DISCUSSION

The discovery of rare, high-risk germline variants has long proven challenging due to their very low frequency, which substantially impacts power to detect significant statistical associations. However, there remains considerable impetus to characterise rare risk variants in DDR genes, especially considering the increasing availability of therapies targeting this pathway. In a candidate gene approach designed to take advantage of large familial PrCa resources, where rare risk variants are expected to be enriched, we examined massively parallel sequencing data from two independent datasets to identify rare, likely deleterious DDR variants. Subsequent analysis of 1,963 individuals from the Australian and *PROGRESS* datasets revealed statistically significant associations between rare variants in *ERCC3* and *BRIP1* and PrCa risk, with *ERCC3* surviving correction for multiple testing. In addition, a variant in *PARP2* was significantly associated with PrCa risk in the Australian dataset alone, while a variant in *MUTYH* was significantly associated with PrCa risk only in the *PROGRESS* dataset.

*ERCC3* encodes one of two ATP-dependent DNA helicases, which are core members of the nucleotide excision repair pathway. The *ERCC3* rs145201970 variant (MAF 0.17%), located in exon 7, causes an amino acid change at position 283 (p.R283C), which is predicted to disrupt the arginine-aspartic acid salt bridge via the inclusion of a more hydrophobic residue. The variant is located within two domains listed by Interpro as likely required for *ERCC3* protein function (31). Topka et al. comprehensively examined germline mutations in the *ERCC2*, 3, 4, and 5 genes in 16,712 patients affected by multiple different cancers (32). Numerous likely pathogenic/pathogenic loss of function (LoF) germline variants were observed in *ERCC3*, with rs145201970 (n=42) representing the second most observed LoF variant in this gene in cancer patients after rs34295337 (n=70)(32). While there are no previous reports describing rs145201970 as a PrCa risk variant, other germline pathogenic/likely pathogenic *ERCC3* variants in PrCa patients were recently reported by Kohaar et al. (33). Additionally, an intronic *ERCC3* variant has been previously associated with increased risk of biochemical recurrence after low-dose-rate prostate brachytherapy, potentially due to reduced mRNA expression in variant carriers (34). In breast cancer, a recurrent truncating mutation has been associated with familial disease (35, 36). *In vitro* studies have demonstrated mutations in *ERCC3* impair DNA repair capability and confer a selective sensitivity to Irofulven, a sesquiterpene that has demonstrated some efficacy in clinical trials for metastatic PrCa (32).

*BRIP1* is a member of the Fanconi Anaemia gene family and functions in the double-strand break repair pathway, interacting closely with *BRCA1*. The rare rs4988345 variant (MAF 0.43%) is in exon 5. As a result of the p.R173C amino acid change, there is a loss of positive charge and a more hydrophobic residue introduced within a helicase ATP-binding domain and a region annotated as a nuclear localization signal. *BRIP1* rs4988345 has been previously identified in a study enriched for familial PrCa but was only observed in a single PrCa case (0.52%)(37). Other rare *BRIP1* variants were detected in five hereditary PrCa cases (MAF <1%)(38), however, no statistical analyses were performed due to their low frequency. A study in breast cancer has linked the rs4988345 variant to disease susceptibility through impairing protein translocation to the nucleus (39). *BRIP1* has been included on screening panels for several clinical trials investigating the response of metastatic PrCa patients with DDR defects to Olaparib, a PARPi (ClinicalTrials.gov Identifier: NCT02987543)(40). A cohort of that study comprised men harbouring mutations in 12 DDR genes, including *BRIP1*, however, only four individuals were identified as carriers of a variant in this gene, below the pre-set threshold for statistical analysis. Evaluation of *BRIP1* has also been included in the Phase 2 TRITON2 trial (ClinicalTrials.gov Identifier: NCT02952534) where one patient with a *BRIP1* variant responded to the PARPi, Rucaparib (41).

*PARP2* is a poly (ADP-ribose) polymerase involved in the base excision repair pathway (BER), and rs200603922 is located in the first exon (p.R15G). This variant (MAF 0.12%) has previously been observed to partially segregate with PrCa in familial cases who tested negative for *BRCA1* and *BRCA2* mutations (42). Although several bioinformatic tools predict the variant allele to be benign (Table 4), the R15G amino acid change introduces a more hydrophobic residue, which may impact protein interactions and the phosphorylation of distal residues. There is one other report of a *PARP2* variant, rs3093926 (MAF 4.2%), segregating in a PrCa pedigree, but the contribution of this variant to PrCa risk remains undetermined (43) and though common, it was not observed in our Australian discovery families. *PARP2* mutations have been associated with breast cancer risk (44), but similar to PrCa, their functional impact remains unclear. However, *PARP2* remains of interest given the ongoing development of PARPi. Though most primarily target PARP1, some, such as Niraparib (45), also affect *PARP2*, which may be relevant when assessing therapeutic PARPi in men with *PARP2* mutations.

*MUTYH* encodes a DNA glycosylase involved in oxidative DDR and the BER pathway. The rs36053993 variant (MAF 0.47%) results in an amino acid change from a neutral residue to a negatively charged, less hydrophobic residue (p.G368D), with this change located in the highly conserved nudix hydrolase domain. The NCBI human variant database, ClinVar, lists this variant as pathogenic/likely pathogenic arising from its association with *MUTYH*- associated polyposis, an autosomal recessive hereditary condition typified by the development of colorectal carcinomas. Kohaar et al. (2022) previously reported the rs36053993 SNP in germline samples from PrCa patients (33). Others have also reported several pathogenic/likely pathogenic variants in *MUTYH*, including a study reporting 1.8% of 1,351 PrCa cases (46), while another reported 2.4% of 3,607 PrCa cases as carrying pathogenic variants in this gene (47). Furthermore, reduced gene and protein expression of *MUTYH* in prostate tumours has been associated with an increase in total somatic mutations, which may result from impaired DDR capacity (48).

In this study, the strategy for filtering and prioritisation of variants was developed to detect highly penetrant, rare DDR gene variants that may contribute to familial PrCa risk. It is notable that rare germline variants predicted to be deleterious have been previously observed in *BRIP1* (n=7), *ERCC3* (n=8), *MUTYH* (n=10), and *PARP2* (n=5) in a cohort of 5,545 non-aggressive and aggressive sporadic PrCa cases (11). While no statistically significant association with aggressive disease risk was observed, due to the very low frequency of these variants, their association with PrCa risk in general was not explored in this case-only cohort (see Supplementary Tables, Darst et al. (11)). Notably, this study examined 5,545 cases and statistically significant associations with aggressive disease were only demonstrated for previously known PrCa risk genes, *BRCA2* and PALB2, with a nominal association seen for *ATM*. This highlights the fact that the innate rarity of DDR gene variants presents a significant challenge for rare variant evaluation, even in larger sporadic case datasets. Our approach was designed to maximise power by seeking to identify rare variants enriched in two large familial PrCa cohorts. However, it is possible that additional rare, disease-associated variants were not detected due to not being present in the Australian WGS discovery cases. It was also necessary to restrict follow-up to only those candidate variants observed in more than one North American family, as the rareness of these variants impacts statistical power to detect associations. However, this may have resulted in risk variants associated with disease in the Australian cohort being missed, e.g., *ATM* rs56128736. Furthermore, instances where prioritised variants were subsequently not found to be associated with PrCa could be attributed to their rarity and, thus, lack of statistical power. Further examination of additional familial and population-based datasets is therefore required to establish the necessary evidence-base to inform clinical decision making. Concurrently, it is worth considering the expansion of candidate gene screening strategies in current clinical trials of PARPi and in current germline testing guidelines for men with a family history of PrCa.

Examining rare variant association with clinicopathological variables presented significant challenges, again due to their rarity, but also due to biases introduced by recruitment strategies. For example, while the aim was to collect all known relatives with PrCa in the Australian and *PROGRESS* familial cohorts, population-based cases with an early age-at-diagnosis were targeted for recruitment to the Australian case-control study.

Carriers of putative pathogenic DDR variants were slightly more frequently observed in the earlier age-at-diagnosis group, and mildly increased in the GS≥8-at-diagnosis group when compared with the population-based PCOR-TAS cohort. However, it is notable that the majority of variant carriers were diagnosed with a GS≤6 (58%) and/or at age ≥65 years (53%), consistent with the findings of Darst et al. (11), where carriers of DNA repair mutations were, on average, diagnosed only ∼1 year younger than non-carriers. Taken together, these findings raise the question as to whether limiting screening for putative genetic DDR variants to very early-onset (<50 years) or only high-grade disease patients is likely to result in a substantial proportion of DDR variant carriers being excluded from testing, and subsequently denied access to effective treatment modalities.

## Conclusions

This study implicates several additional DDR genes as contributors to inherited genetic risk of PrCa. The existing evidence that rare DDR gene variants are associated with aggressive disease and the growing use of cancer therapies targeting this pathway highlights the potential significance of these findings. However, this study raises the concern that confining genetic screening to only those patients with early onset and/or high-grade PrCa may result in a missed opportunity for some men to receive effective, targeted gene-based therapies.

## Supporting information

Supplementary Results Tables

## Data Availability

The data underlying this study are available in the article and its online supplementary material, or from the corresponding author on reasonable request. The genome and exome sequencing data underlying this article cannot be shared publicly for ethical/privacy reasons.

## Abbreviations

BER: base excision repair
CADD: Combined Annotation-Dependent Depletion
DDR: DNA damage repair
FHCC: Fred Hutchinson Cancer Centre
GS: Gleason Score
MAF: minor allele frequency
M_QLS_: modified quasi-likelihood score
NFE: non-Finnish Europeans
PARPi: poly (ADP-ribose) polymerase inhibitors
PCOR-TAS: Prostate Cancer Outcomes Registry – Tasmania
PrCa: prostate cancer
PSA: prostate specific antigen
TCR: Tasmanian Cancer Registry
WES: whole-exome sequencing
WGS: whole-genome sequencing

## ADDITIONAL INFORMATION

## Acknowledgements

We are greatly indebted to the participants of our prostate cancer studies, the Tasmanian Cancer Registry staff, Tasmanian urologists, pathologists, and the wider Tasmanian clinical and research community for their ongoing support. We also thank the individuals participating in the FHCRC *PROGRESS* study.

## Authors’ Contributions

Conceptualization – LMF, JLD; Data curation – GRF, JRM, SEL, KR; Formal Analysis - GRF; Funding acquisition – LMF, JLD; Investigation – GRF, JRM, SEL, LMF, JLD; Methodology – GRF, JRM, SEL, KR, LMF, JLD; Project administration – JLS, LMF, JLD; Resources – JRM, AB, JLS, EAO, LMF, JLD; Supervision – LMF, JLD; Validation – GRF, JRM, JLS, EAO; Writing – original draft – GRF, JRM, LMF, JLD; Writing – review & editing - all authors.

## Ethics Approval and Consent to Participate

Ethics approval for this study was obtained from the Human Research Ethics Committee Tasmania, Australia (H0017040). Written informed consent was gained for all participants. PCOR-TAS participants consented to their data being used for ethically approved research and ethics approval to use these data was obtained from the Tasmanian Health and Medical Human Research Ethics Committee, Australia (H0017095). The *PROGRESS* study was approved by the FHCRC’s Institutional Review Board and informed consent was obtained from all study participants. All participants provided consent for inclusion of their data in publications.

## Data Availability

The data that supports the findings of this study are available in the article and its online supplementary material, or from the corresponding author upon reasonable request. The genome and exome sequencing data are not publicly available due to privacy or ethical restrictions.

