## Supplementary Methods for "Germline sequencing of DNA-damage-repair genes in two hereditary prostate cancer cohorts reveals new disease risk-associated gene variants"

**Supplementary Method 1: Nucleic Acid Extractions and Whole Genome Sequencing**

Germline DNA was extracted from either 10mL of peripheral blood using the Nucleon BACC3 Kit (GE Healthcare) or from 4mL of saliva using the Oragene OG-500 DNA collection tubes (DNA Genotek), following the manufacturers’ directions. All DNA samples were quantified using a Nanodrop 8000 UV-vis spectrophotometer (Thermofisher Scientific) and normalised for whole-genome sequencing (WGS) and/or TaqMan™ Genotyping.

**Supplementary Method 2: Statistical Association Analysis**

Modified Quasi-Likelihood Score (M_QLS_) analysis permits the combined analysis of genotype data from both familial and case-control samples, taking relatedness into consideration. It also distinguishes between unaffected and uncertain phenotypes, incorporating both into the analyses. M_QLS_ uses variance components to examine the significance of association for related individuals, and when the disease status is known for first-degree relatives of cases, M_QLS_ obtains additional power by giving increased weighting to those individuals with closely related disease-carrying relatives. The use of this approach maximises power but does not inflate type 1 error (1, 2).

**Supplementary Method 3: Bioinformatic Predictions**

The predicted structural and functional effects of candidate risk variants was evaluated using the online analysis server, HOPE (3). Protein sequences and amino acid information was sourced via the UCSC genome browser (4). Additional bioinformatic tools, including DANN (5), Sift (6), PROVEAN (7), PolyPhen (8), Mutation Taster (9), Mutation Assessor (10), FATHMM (11), and Protein Predict (12) were used to appraise the predicted pathogenicity of candidate variants.
