## Supplementary Results Tables for "Germline sequencing of DNA-damage-repair genes in two hereditary prostate cancer cohorts reveals new disease risk-associated gene variants"

**Supplementary Table S1: Summary of individuals from the Australian discovery cohort with WGS data**

| **Family** | **PrCa Cases** | **PrCa Unaffected Relatives** | **Total** |
| --- | --- | --- | --- |
| PC2 | 7 | 2 | 9 |
| PC3 | 5 | 0 | 5 |
| PC4 | 5 | 0 | 5 |
| PC9 | 8 | 1 | 9 |
| PC12 | 2 | 1 | 3 |
| PC18 | 3 | 0 | 3 |
| PC22 | 9 | 3 | 12 |
| PC72 | 4 | 4 | 8 |
| Total | 43 | 11 | 54 |

**Supplementary Table S2: Genes extracted from WGS data**

| **Gene** | **Chromosome** | **Start Position (bp*)** | **End Position (bp*)** |
| --- | --- | --- | --- |
| *ATM* | 11 | 108092559 | 108240826 |
| *BARD1* | 2 | 215592275 | 215675428 |
| *BLM* | 15 | 91259579 | 91359686 |
| *BRCA1* | 17 | 41195312 | 41278500 |
| *BRCA2* | 13 | 32888617 | 32974809 |
| *BRIP1* | 17 | 59755547 | 59941920 |
| *CDH1* | 16 | 68770195 | 68870444 |
| *CDKN1B* | 12 | 12869302 | 12876305 |
| *CDKN2A* | 9 | 21966751 | 21976132 |
| *CHEK2* | 22 | 29082731 | 29138822 |
| *ERCC2* | 19 | 45853649 | 45874845 |
| *ERCC3* | 2 | 128013866 | 128052752 |
| *GEN1* | 2 | 17934177 | 17967632 |
| *HOXB13* | 17 | 46801127 | 46807111 |
| *MLH1* | 3 | 37033841 | 37093337 |
| *MRE11A* | 11 | 94149469 | 94228040 |
| *MSH2* | 2 | 47629206 | 47711367 |
| *MSH5* | 6 | 31706725 | 31731945 |
| *MSH6* | 2 | 48009221 | 48033092 |
| *MUTYH* | 1 | 45793914 | 45807142 |
| *NBN* | 8 | 90944564 | 90997899 |
| *PALB2* | 16 | 23613483 | 23653678 |
| *PARP2* | 14 | 20810773 | 20827063 |
| *PMS2* | 7 | 6011870 | 6049737 |
| *POLD1* | 19 | 50886580 | 50922275 |
| *POLE* | 12 | 133199348 | 133264945 |
| *PTEN* | 10 | 89622195 | 89729532 |
| *RAD50* | 5 | 131891616 | 131981313 |
| *RAD51C* | 17 | 56768963 | 56812692 |
| *RAD51D* | 17 | 33425811 | 33434500 |
| *RECQL4* | 8 | 145735667 | 145744210 |
| *SLX4* | 16 | 3630184 | 3662585 |
| *STK11* | 19 | 1204798 | 1229434 |
| *TP53* | 17 | 7570720 | 7591868 |
| *XPC* | 3 | 14185648 | 14221172 |

*Gene positions according to hg19.

**Supplementary Table S3: Primers used for Sanger sequencing**

| **Gene** | **Variant** | **Forward Primer (5'-3')** | **Reverse Primer (5'-3')** | **Amplicon Size (bp)** |
| --- | --- | --- | --- | --- |
| *ATM* | rs55801750 | AAAGGTTCAGCGAGAGCTGG | GATTCCTGACATCAAGGGGCT | 273 |
| *ATM* | rs55982963 | TCTAAATGAAAGAATGGCAGTAGGT | CCTAGTTTCCGTGTTTCTCTGC | 250 |
| *ATM* | rs767507047 | GATGTTTGTTCCCCTCCCCC | TGAAAAACTGACAACAGGACCTT | 305 |
| *ATM* | rs56128736 | TGAAGATACCAGATCCTTGGAGA | AGGTTTGGGGGTAGACAAATGA | 285 |
| *BARD1* | rs3738888 | AGCTGTTGAAAGGGCAGAAGT | TGCCATGAAGAAGAAAAACCACT | 293 |
| *BRCA1* | rs28897673 | ACCAAGGAAGGATTTTCGGGT | CACAACAAAGAGCATACATAGGGT | 243 |
| *BRCA2* | rs786202915 | TTCTGATGTTCCTGTGAAAACAAA | GTGATTGGCAACACGAAAGGT | 279 |
| *BRCA2* | rs55639415 | ACTTCTGTGAGTCAGACTTCATT | TCTTCAATACTGGCTCAATACCAG | 298 |
| *BRCA2* | rs56403624 | GTACCGTCTTTGGCCTGTGA | TTGCCTGCTTTACTGCAAGA | 301 |
| *BRCA2* | rs28897727 | TGGCCAGTTTATGAAGGAGGG | GGAAAAGTTATGCAATTCTTCTGGT | 277 |
| *BRIP1* | rs4988345 | TGGCATTAATACATACTTTCTGTGG | GTTGTAATGAGGTGCTTATTTGCAT | 310 |
| *ERCC2* | rs142568756 | AATGAAGCTGACATAGCGGTG | CTAAGACAGAGAAGGGAGGAGGA | 257 |
| *ERCC3* | rs145201970 | CAGCTGTGGGCTTTAGGTCA | CCGGTTGTTGACTGAGCAAG | 264 |
| *MRE11* | rs777373591 | AGGCATGCTTTCCACAGACA | TGCAGTTTGCCTATGATTGCATTA | 261 |
| *MSH6* | rs142254875 | ACTTAGGCTGATAAAACCCCCAAA | GCTCCTCTTCCTCACAGCCTA | 251 |
| *MUTYH* | rs36053993 | CATCCTTGGCTATTCCGCTG | ACCTGGATACTGGGCGTG | 268 |
| *PARP2* | rs200603922 | CCTGTCCCTCACAGCCATCTTC | AGGTTATAGGGAGCTGGAAGGG | 264 |
| *PMS2* | rs1554304601 | TGGCAGCGAGACAAAACAGA | TCCTTACTTTACACTCTCTTTCAGC | 265 |
| *POLE* | chr12:133219216 | GTGCTCACCTGCTCATCTCG | GGTCCACCCAGGTCTTTTCT | 272 |
| *POLE* | rs36120395 | CAACGCAGCCCAGTAAGAAC | TTTGAACTTGCCCCCATTGC | 254 |
| *POLE* | rs41561818 | GGCGGGTTTCTTTCCTCCAT | GACTCCGAATAGCGTGTGCT | 299 |
| *PTEN* | rs773513402 | GGTAAGAAACACAGCAACAATGAC | GCAGCACATGAAGCATCCAC | 254 |
| *PTEN* | rs587779989 | TGGGACGCGACTGCG | GGCTGCACGGTTAGAAAAGAC | 251 |
| *RAD51C* | rs61758784 | TATTCTTGGGGGTGGAGTGC | TGTTTCTTTTGCAAATTGTACTGCA | 310 |
| *RECQL4* | rs780723602 | CCTGATTCTCCAACCTCGTCT | TGAGCGGGCACTCCCAATA | 305 |

**Supplementary Table S4: TaqMan**™ **genotyping assay information**

| **Gene** | **Variant** | **TaqMan™ genotyping assay (ThermoFisher Scientific)** |
| --- | --- | --- |
| *BARD1* | rs3738888 | C__27471488_10 |
| *BRCA2* | rs28897727 | C__11711256_20 |
| *BRIP1* | rs4988345 | C__2649849_20 |
| *ERCC3* | rs145201970 | C_168471925_10 |
| *MUTYH* | rs36053993 | C__27860252_10 |
| *PARP2* | rs200603922 | C_190044127_10 |

**Supplementary Table S5: Prostate cancer status of variant carriers**

| Gene | Variant | Tasmanian Familial Prostate Cancer Study (Discovery Families) | | | | | *PROGRESS* | | Total |
| --- | --- | --- | --- | --- | --- | --- | --- | --- | --- |
|  |  | **Familial Cases** | **Unaffected Male Relatives** | **Female Relatives** | **Sporadic Cases** | **Sporadic Controls** | **Familial Cases** | **Unaffected Male Relatives** |  |
| *BARD1* | rs3738888 | 8 (2) | 5 (3) | 4 (0) | 10 | 4 | 4 | 0 | 35 |
| *BRCA2* | rs28897727 | 6 (3) | 7 (5) | 4 (1) | 7 | 0 | 4 | 1 | 29 |
| *BRIP1* | rs4988345 | 7 (2) | 1 (1) | 6 (2) | 9 | 2 | 6 | 0 | 31 |
| *ERCC3* | rs145201970 | 5 (2) | 4 (4) | 1 (1) | 3 | 3 | 5 | 0 | 21 |
| *MUTYH* | rs36053993 | 2 (2) | 5 (2) | 7 (1) | 4 | 5 | 9 | 0 | 32 |
| *PARP2* | rs200603922 | 6 (4) | 3 (1) | 1 (1) | 4 | 0 | 2 | 0 | 16 |
